# Signal from the Noise: A Mixed Methods Process Mining Approach to Evaluate Care Pathways

**DOI:** 10.1101/2021.11.08.21266066

**Authors:** Morteza Noshad, Christian C. Rose, Jonathan H. Chen

**Author notes:** Corresponding Author: Christian Rose, MD, 900 Welch Road, Suite 350, Palo Alto, CA 94304. Both authors contributed equally (Morteza Noshad, Christian C. Rose).

## Abstract

**Objective:** Mapping real-world practice patterns vs. deviations from intended guidelines and protocols is necessary to identify and improve the quality of care for emergent medical conditions like stroke. We propose a process mining algorithm applied to Electronic Health Record (EHR) event log data as a unique opportunity to more easily identify and compare real-world care processes.

**Materials:** Data was obtained from the event log of a major EHR vendor (Epic) for Stanford Health Care Hospital patients aged 18 years and older presenting to the ED from January 1, 2010 through December 31, 2018 and receiving tPA within 4.5 hours of presentation. Our algorithm was built using the Python programming language.

**Methods:** An unsupervised process-mining algorithm was developed and used to create a process map for our cohort. This map was then used to identify the most common path as well as individual and average conformity to this path across all encounters.

**Results:** Our automatically generated process mining graph, specifically its most common path, mimicked our institution’s recommended “code stroke” clinical pathway. The average conformity score for our cohort was 0.36 with a range from high of 0.64 and low of 0.20.

**Discussion:** This method allows for greater detail into common process measures to be more easily illustrated to evaluate the quality of care at a given institution. It may be extended to other, similarly well-defined processes or those which currently lack standardized clinical pathways.

**Conclusion:** Our mixed methods approach represents an essential data analysis step to improve complex care processes by automatically generating a qualitative and quantitative process measures from existing event log data which can then be used to target quality improvement initiatives.

## INTRODUCTION

Many life-threatening emergent medical conditions require time-sensitive progression through multiple points of evaluation and management in order to deliver definitive, life-saving interventions. In the case of an acute ischemic stroke, for example, blood samples must be collected by a nurse, evaluation must be performed by a physician, the CT scanner must be prepared by a technician and a pharmacist must prepare critical “clot-busting” medication like tissue plasminogen activator (tPA) for delivery as soon as the diagnosis is confirmed. Each step may rely on another and variation may result from institutional and contextual differences in the care environment[1]. Evaluating and improving care processes for conditions such as these underlies the vision of a learning healthcare system[2].

Before attempting to improve such processes, however, we must understand the status quo practices[3]. Only then can we begin to measure its variation and identify discrepancies between common versus intended practices to drive improvement initiatives. However, given the complexity of most real-world healthcare processes, it is difficult to obtain granular detail about how they actually occur in practice[4]. Defining clinical care processes currently tends to rely on recall and expert opinion rather than direct observation[5]. When they are directly observed, they tend to occur in-person, which can be time-consuming and labor-intensive. Video surveillance may also be utilized, but managing emergent conditions often takes place in multiple simultaneous settings (ie. the ED, hallways, or the radiology suite), thus limiting their generalizability to those actions that occur at the bedside.

Process mining, the method of determining the order of events from a log, may help to address these issues[6]. All that is required is time-stamped data, that is often generated at scale through passive data collection in electronic (health) record (EHR) systems. Process mining can help organizations easily capture workflow information from enterprise systems and provides detailed, data-driven insights about how key processes are being performed.

In the clinical setting, process mining has been utilized for various purposes[7, 8, 9]. Most of these studies evaluate how closely a process matches recommended guidelines (ie. appropriate triage),[10, 11] while some assess variations in practice for poorly defined workflows (ie. what is an average day for a general surgeon)[12]. This methodology has been recommended as particularly useful in the ED setting, where practice and processes are highly variable and noisy, but limited existing investigation work has been done in this setting[7, 13]. Cho et. al. proposed several common ED process measures which could be evaluated through this method, yet recommendations for which data to use and what events to measure remains unclear[14].

Furthermore, even when an ED process has been evaluated, as in the case of acute stroke, the methodologies thus far have not evaluated the granular details of diagnosis and treatment (e.g., medication ordering, administration and CT scanner availability) but rather the department-level identifiers like wait times and level of service[13], which limits their potential utility in affecting performance improvements at the point of care. Finally, while there are many different process mining algorithms and software packages available for use, there is little consensus on which to use and for what purposes, few are able to tolerate the complexity and volume of data from raw healthcare event log data sets, and none employ unsupervised machine learning strategies to clean and present the most relevant data[7].

In this exploratory data analysis, we developed a process mining method to automatically analyze the event log from a common EHR vendor, Epic, at an academic medical center (Stanford Health Care) to build process maps for tPA management of acute stroke care solely from the event log data. We evaluated our model’s process maps to our institutional stroke guidelines to validate our proof of concept. Furthermore, we extended the visualizations and insights such maps can offer by providing additional summary views and statistics to illuminate variations in care practices.

## METHODS

Process mining is based on a set of simple rules to create a graphical representation of actions. Actions are acquired from the event log of a particular outcome of interest (ie. tPA administration for stroke). Process mining results in a graph where events are represented by nodes, edges represent subsequent events, and edge labels show the probability of those two events occurring in that order. The nodes of the graph are time-ordered earliest to latest (top to bottom).This graph is then used to determine the most common pathway (MCP). The MCP is then used to calculate the conformity of an individual encounter as well as the average conformity of all encounters to it.

### Cohort Development

First, a cohort of patients and data from their hospital encounters must be created based on inclusion criteria for a particular process or outcome of interest. We investigated patients receiving tPA treatment for acute stroke. As such,our cohort consisted of Stanford Health Care Hospital patients aged 18 years and older who presented to the ED from January 1, 2010 through December 31, 2018 and received tPA within 4.5 hours of presentation - the standard window for safe tPA administration recommended by the American Stroke Association (ASA). Given this cohort, data may be extracted or filtered from the EHR event log for their corresponding clinical encounters.

### Data Preparation

Given the volume of clinical event log data is orders of magnitude greater than clinical data alone, feature selection is critical before creating a process map.

The event log for a unique patient encounter includes several columns such as “user ID”, “time”, “event name”, etc. The “user ID” specifies the unique ID for a provider (Table 1). The “time” column indicates the relative times of events. These relative times are the difference between the patients’ admission time and the time of the associated event. Negative times indicate events which have happened even before the admission time (ie. activation of stroke protocol from the field before patient arrival in the ED). We included data from the time of an encounter beginning through the administration of tPA.

**Table 1:** The event log of a simulated patient.

| user_id | time | event_type | event_name | event_category | user_type |
| --- | --- | --- | --- | --- | --- |
| 1 | 7 | Initiate Procedure | ISTAT INR AND PROTIME | Coagulation Studies | Emergency Resident |
| 1 | 7 | Lab Result | ISTAT INR AND PROTIME | Coagulation Studies | Emergency Resident |
| 1 | 9 | Initiate Procedure | ISTAT CREATININE | Basic Labs | ED Nurse |
| 1 | 9 | Lab Result | ISTAT CREATININE | Basic Labs | ED Physician |
| 1 | 15 | Initiate Procedure | CT HEAD | CT Head | ED Physician |
| 1 | 42 | Order Medication | ALTEPLASE 100 MG IV SOLR | tPA | ED Physician |
| 1 | 42 | Lab Result | PROTHROMBIN TIME | Coagulation Studies | Emergency Resident |
| 1 | 42 | Lab Result | PTT PARTIAL THROMBOPLASTIN TIME | Coagulation Studies | ED Physician |
| 1 | 43 | Lab Result | ECG 12-LEAD | ECG 12-LEAD | Emergency Resident |
| 1 | 45 | Initiate Procedure | XR CHEST 1V | CXR | ED Nurse |
| 1 | 45 | Initiate Procedure | PROTHROMBIN TIME | Coagulation Studies | Emergency Resident |
| 1 | 45 | Initiate Procedure | FOLEY RETENTION CATHETER | FOLEY RETENTION CATHETER | ED Physician |
| 1 | 45 | Initiate Procedure | PTT PARTIAL THROMBOPLASTIN TIME | Coagulation Studies | ED Nurse |
| 1 | 45 | Initiate Procedure | FOLEY RETENTION CATHETER | FOLEY RETENTION CATHETER | Emergency Resident |
| 1 | 45 | Initiate Procedure | ECG 12-LEAD | ECG 12-LEAD | ED Physician |
| 1 | 45 | Initiate Procedure | PATIENT ON ACTIVE PROTOCOL | PATIENT ON ACTIVE PROTOCOL | Emergency Resident |
| 1 | 47 | Medication Given | ALTEPLASE 100 MG IV SOLR | tPA | ED Nurse |

The “event name” column contains specific names of the events (i.e. order name or result that was viewed). The event log table structure can contain additional columns to assign more information or labels for each event. In our example these event label columns are “event category”, “event type” and “user type.”

### Process Mining Algorithm

Our implementation of process mining utilizes an unsupervised learning method to create the subsequent graph.

#### Automatic Selection of Events

Many unique event types can occur in a clinical process, most of which are captured in the clinical EHR data. Figure 2 illustrates the timing distribution of some of these most common events for our cohort. The corresponding event log data typically includes even more unique events of varying prevalence. Our proposed approach for selecting events that should appear as nodes in a process mining graph involves selecting the top *n* events that happen most frequently among all patients from the event log data. A larger *n* would result in a graph with more nodes, which captures a more granular set of events in the process.

#### Graph Construction

There is a relatively normal distribution in timing of common events as seen in Figure 1. However, the temporal relationship of each order to its pro- and pre-ceding events cannot be determined from the summary timing data alone.Instead, all must be mapped from beginning to end for each patient encounter. Mapping relies on creating a graph of nodes and their related edges.

**Figure 1:**
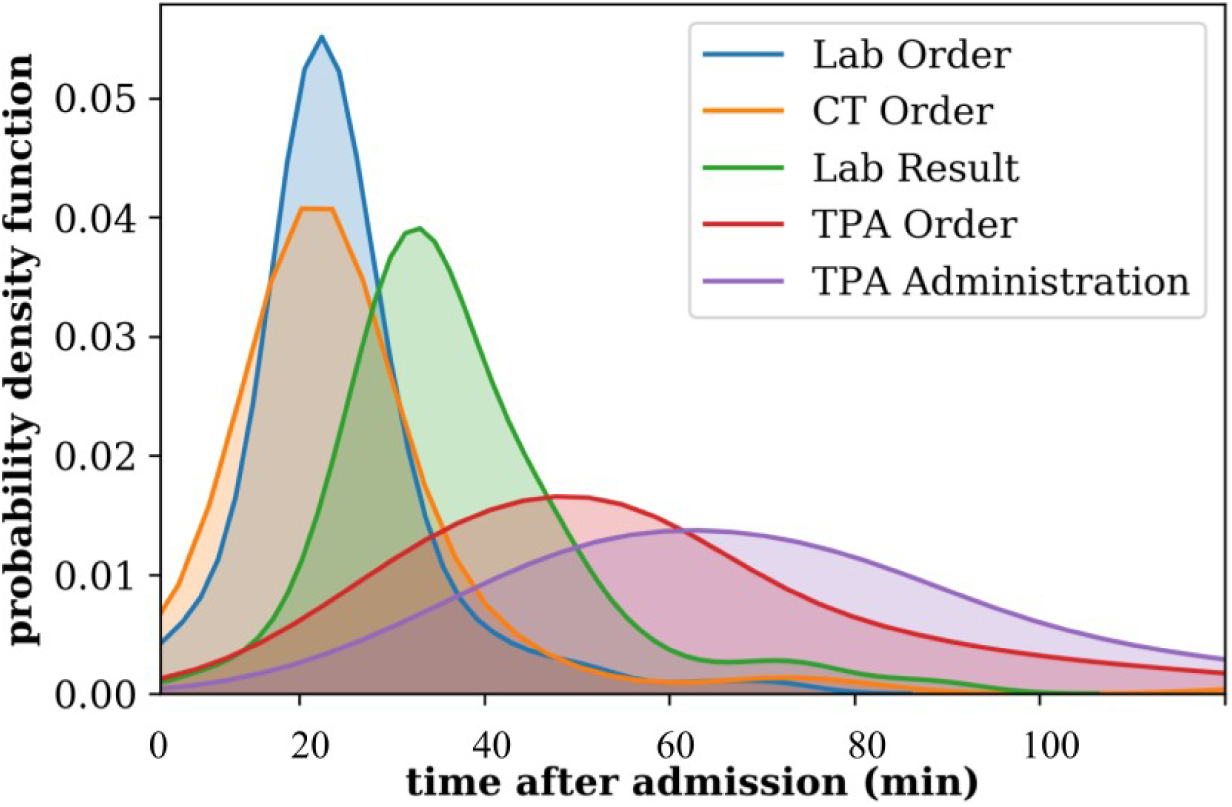
Histogram of the major events for stroke patients in the ED.

**Figure 2:**
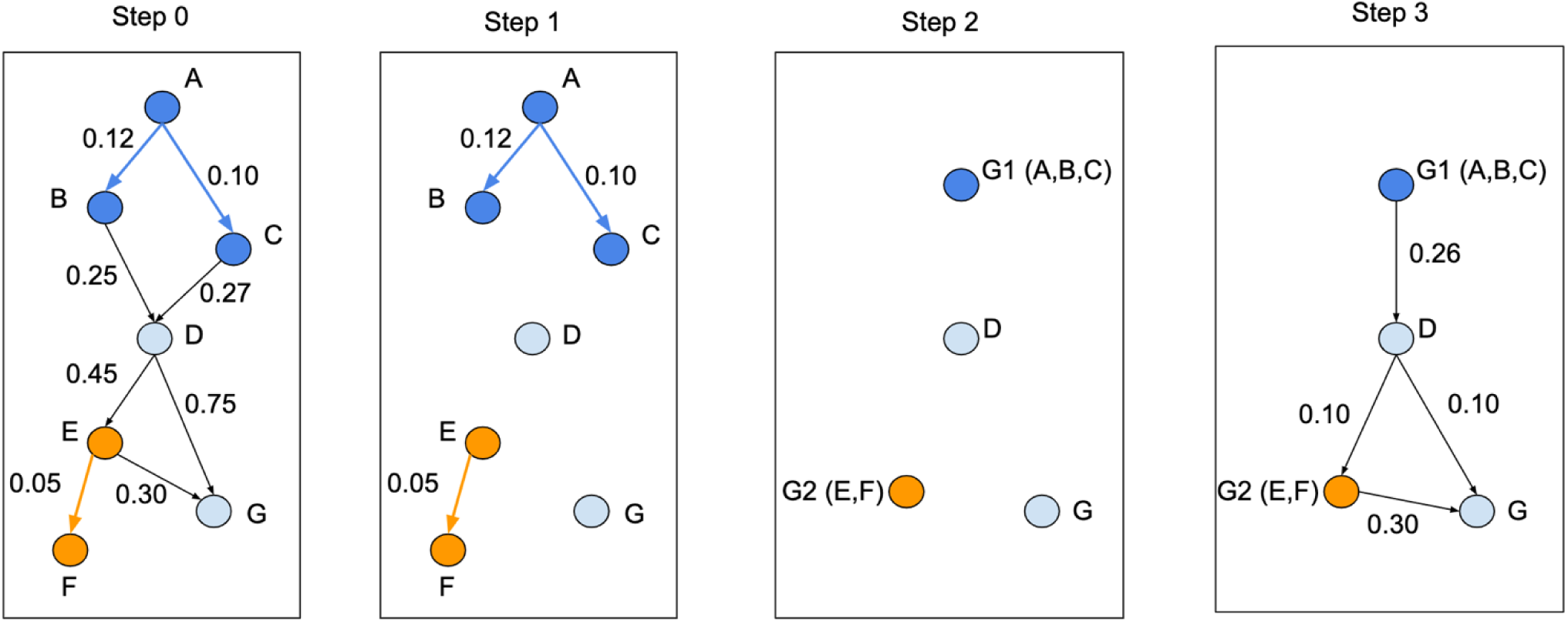
Node aggregation example. Step 0: the original process mining graph, Step 1: filter the edges, Step 3: node aggregation, Step 4: edge aggregation.

### Nodes

The nodes of the graph are the unique events from the event-log data. The node names can be defined based on any set of the event labels (associated with different columns in the event log data (Table 1)).

### Edges

The edges of the process mining graph connect the pairs of events that happen sequentially in the event-log data. It is of course possible for many variations in the ordering of these events. Without applying a filter the output graph the resulting graph might appear complex and difficult to interpret. In order to control the number of the edges we define two types of weights for each edge:

#### Probability weight

The probability weight is proportional the number of occurrences of particular nodes pairs. The weights are then normalized.

#### Lag weight

The lag weight for each edges shows the relative time between the corresponding end-nodes.

As a direct pruning measure, less frequent edges with a probability weight below specified thresholds may be removed from the graph.

##### Node Aggregation

Limiting the number of edges is one way to reduce noise and improve the readability of the process mining graph. Many separate events often occur simultaneously or with an extremely small time lag (ie. using an order set to place many simultaneous orders). In these situations aggregation of the events into a single node better represents the action associated with this process. Detecting the set of nodes which need aggregation is based on a graph-theoretical concept of graph clustering^14^. Node aggregation is based on the time-lag between the events as well as the event labeling - to avoid unique but simultaneous events from being aggregated. We refer to the new graph with aggregated nodes as the *summarized* graph.

Aggregation occurs as follows:

1. **Filter edges:** Filter out the edges with time-lag weights greater than a predefined threshold.
2. **Aggregate Nodes:** Identify nodes that belong to the same cluster and aggregate them into single nodes.
  a. **Identify the connected components:** After removing the edges with larger time-lags, the resulting graph has several connected components (clusters). Larger weight thresholds leads to larger clusters and more node aggregations.
  b. **Exclude the nodes from the minority types:** The nodes with different types (e.g. with different categories, users, etc.) are not usually associated with similar sets of events. Thus, nodes with different types from each connected node set are excluded.
  c. The nodes within each cluster are then grouped into a <u>single node</u> representing the whole cluster.
3. **Aggregate Edges:** The final step is to aggregate the edges connected to the aggregated nodes. This includes two types of edges:

#### Internal edges

An edge that connects two nodes within a cluster is called an internal node. These edges disappear in the summarized graph.

#### External edges

An edge that connects two nodes from a cluster to another node outside the cluster is called an external edge. In the summarized graph, all of the edges between two clusters would be aggregated into a single edge. The lag-weight and probability weights of the new edge are respectively equal to the average lag-weights and probability weights of the original edges between those clusters. Node aggregation is demonstrated in Figure 2.

##### Most Common Path

One of the main outcomes of a process mining graph is to show the many common pathways among all users. However,it may be useful to identify the MOST common path given many possible routes. The probability weights indicate which order of events most commonly happened among all patients. Based on the probability weights of edges in the process mining graph, we define the MCP as the path from the start node (admission) to the end node (tPA given) with the greatest probability of occurring.

### Pathway Conformity Scores

While the primary application of the proposed process mining graph is to *qualitatively* represent common pathways for a process of interest, it may also be helpful to *quantitatively* measure the conformance of the pathways across patients or encounters for a given outcome of interest. This provides a measure of how similar an individual or cohort of encounters is with the MCP. The conformity score allows for a summary statistic of the totality of individual events from an encounter to be compared against all other encounters for that outcome of interest. Here, we define the pathway conformity score for a particular patient encounter as well as the average conformity score for all encounters of a particular outcome.

#### Patient-Level Conformity Score

We define the conformity score for an individual patient encounter *P* using the probability weights of the edges as follows:

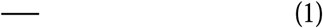

where *P* represents a path for a patient, *E*_*P*_ is the set of all of the edges for the patient *P*, and *p*_*e*_ are the probability weights for the edge *e*. The score is normalized and represents the weighting of a pathway against all other possibilities. For example, if there were three possible unique pathways to take based on three unique encounters, the conformity score for any one pathway would be 1*/*3 (0.33). A higher conformity score means the encounter is more similar to the most common path, whereas lower conformity scores are associated with encounters that proceeded through a much different series of events.

#### Average Conformity Score

It may be helpful to determine how variable all encounters are to the MCP. Thus, we define the average conformity score, which measures the average conformity across all encounters as follows:

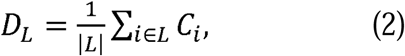

where *L* is the set of all patients. Higher scores imply that the processes are very consistent across different cases - where each encounter has a high conformity with any other on average. If the number is very low, the process is highly variable, or inconsistent, across individual encounters - meaning each encounter has little conformity with any other.

## RESULTS

### Process Map

Using the above-described method, we created a process map for acute stroke management with tPA. There were a total of 269 patients in our above-described cohort, with 140 identified as female and 129 as male gender. The majority of the patients (80%, 214*/*269) were aged 60 or older. The average measured time to tPA is 54 min from admission and the standard deviation is 17 min.

Our process map (Figure 3) showed that admission to the Emergency Department was most often the first clinical event in the stroke care process occurring at time zero (the reference time of our data set). Ordering of head CT occurred on average in the first minute of the encounter, followed by coagulation studies, basic labs, ECG, and Chest X-Ray, respectively. These orders were then followed by their results: basic lab studies, ECG and coagulation studies,respectively. Based on the node aggregation feature of our process mining method, 10 events related to the initial order procedures such as CT Head Perfusion With Contrast, Prothrombin Time, CT Head, etc. are aggregated into a single node. Clinically all of these events correspond to the same checklist. The next most common event was the identification of the start of the Head CT an average of 18 minutes later. The result of the head CT followed, concluding the primary diagnostic testing of these acute stroke patients.

**Figure 3:**
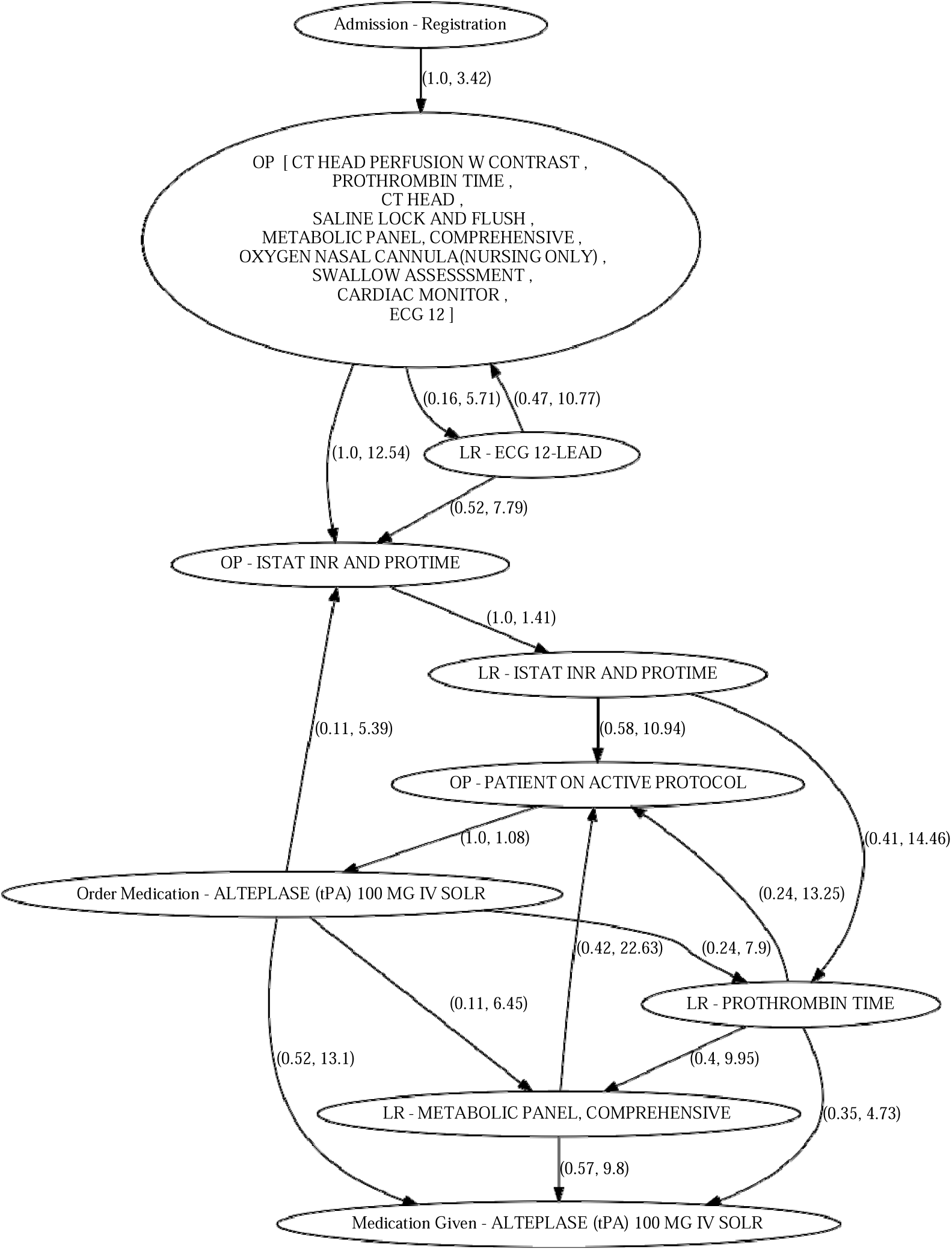
A sample process mining graph of the tPA-treated stroke patients. The following abbreviations are used in the node names: ‘OP’ for ‘Order for Procedure’, ‘LR’ for ‘Lab Result’, ‘ISTAT’ for ‘iSTAT point-of-care testing’.Every edge is labeled by a two-component weight; probability of the sequence (the first weight component) and the lag between events (the second weight component).

Subsequent placement on an active stroke/tPA protocol (including neurologic exam and blood pressure monitoring) followed at about 38 minutes from admission, and tPA was ordered on an average at 39 minutes from admission. tPA was then given on average at about 64 minutes into the encounter.

Evaluation of our cohort showed a subset of acute stroke cases were managed in under 25 minutes (See Figure 3). A process map was also created for these cases to compare to overall results and identify possible differences in management.

For those cases which were managed in under 25 minutes. Head CT’s for these patients were begun at about six minutes from arrival. By minute seven, tPA was ordered and they were started on a protocol. tPA was subsequently given to these patients at an average of the 17th minute. Although these cases progressed faster through the acute stroke pathway, they showed a similar order of events to other cases.

### Conformance

As expected, some encounters varied greatly from the MCP to provide definitive treatment. Some differences resulted from delays in ordering CT scans until after lab results or waiting for radiology result before medication administration.

The average conformity score for our cohort was measured at 0.36. The scores for the patients with the highest and lowest conformity scores are respectively 0.64 and 0.20. Figure 4 represents the process for the patient with the highest conformity score (also referred to as most common path).

**Figure 4:**
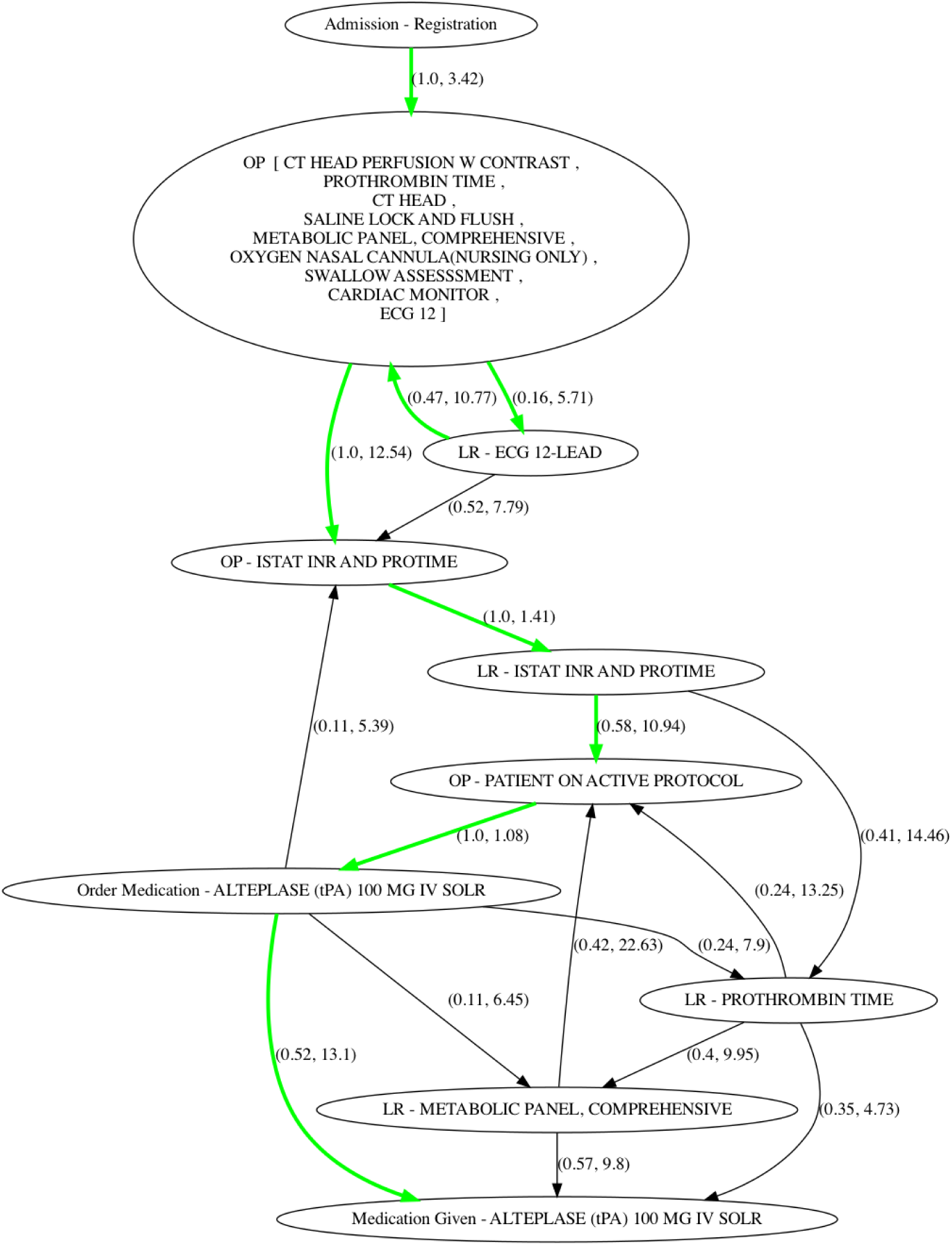
The process mining graph of the stroke patients with green path representing MCP; the path for the patient with the highest conformity score (0.64). The average conformity score for this cohort was measured as 0.36, which is within the expected range. Based on the definition of the average conformity score this means that each node has an average probability of about 0.36, which is in agreement with the fact that every node is connected to about 3 subsequent nodes for this graph.

One of the most obvious examples of deviation from the average pathway lead to the definition of a specific patient cohort subgroup. Given that a subset of all tPA-treated stroke patients were managed from out-of-hospital stroke activation, their ordering and timing of events (as described above) tended to deviate the most from the average tPA process.

## DISCUSSION

Process mining offers a unique opportunity for easily producing human-interpretable results through unsupervised machine learning methods on commonly available data. Our model produced both visual and statistical results that were consistent with the expected order of key interventions in the management of acute stroke as described by our hospital’s “Code Stroke” policy. Ordering and results of lab studies and CT scans as well as ordering and delivery of tPA occurred in the expected order and within the expected window for our institution, providing confidence in the accuracy of this methodology.

This methodology has many possible use cases. Here, we used it to elucidate deeper insight into a common quality measures, which tend to offer limited granular insight into real-world processes. Currently, measurement and reporting of the quality of acute treatment for stroke care at most institutions relies on a single number: the time-to-tPA.[16] However, as we have seen here, this number is made up of an amalgamation of multiple interactions, orders, results and assessments. But there are many other quality measures like “door-to-needle time” or “time to antibiotic administration” which have similar processes and are broadly used throughout medicine. Our process mining method offers a simple way for hospitals to illuminate these events and their impact on care. It may be applied to the events surrounding patient care or any or any common processes - like lab throughput or patient preparation for surgery.

More than just the time and order of events, however, it may also help an institution measure the breadth, or variability, of these pathways. There are generally “common” pathways through a particular process which can be used to determine the status-quo. It may then be useful to determine if that process follows an expected path or deviates from the norm. At our institution, the MCP followed the recommended guidelines. But what if it did not? What, then, is an acceptable amount of deviation from the guidelines? Traditionally, this has not been a quantifiable result, but rather a binary “yes” or “no” answer. Here, we showed that process mining can be used to measure the conformance to recommendations of an entire cohort or between individual encounters. This offers a poignant quality improvement avenue in the age of the EHR.

Deviations and delays in care, or highly variable pathways, may illuminate the need for high-yield, evidence-based interventions. They may be due to lack of knowledge of the guidelines, meaning an opportunity for education, or there may be inherent systematic issues that prevent the medical team from fulfilling them consistently or efficiently, like how labs are ordered or how the CT scanner is made available. Deviations in care might result from patients presenting with ambiguous complaints, making a detailed history and physical more time-consuming. Or a provider may have anchored to a different diagnosis before stroke became obvious – thus presenting as a completely different path to ultimate diagnosis and management. Many stroke patients do not present as clearly as having frank, lateralizing symptoms with a facial droop, but rather with mental status changes that can obfuscate the ultimate diagnosis. This is the case for many medical conditions which require timely, expedited care like “atypical chest pain” or “altered mental status” as a sign of severe sepsis. While all of these cases may seem to result in delayed care, the series of steps along the diagnostic and management pathway might at least be evaluated and inferred, allowing institutions to determine if delays in care result from a changing clinical scenario versus a simple lack of following institutional practice guidelines.

Conversely, patterns in conformity or deviation may serve to illuminate leading-edge care pathways. We found that a set of cases which had particularly low conformity scores may have actually represented higher quality care. Patients who received rapid (< 25 minute) management of their strokes occurred with clustered orders and narrow time windows, resulting in low conformity scores with the MCP, but overall improved care by the quality metric of choice. While they deviated from the average pathway, these cases represent the best possible care which can be provided. Analysis of these cases revealed that they likely resulted from out-of-hospital stroke activations - given that the first orders were placed minutes before the patient was registered in the ED. This allows one to infer variation in care processes simply from the automatically-recorded data which can then be quantified to determine the relative impact that variation has on timing and subsequent process measures. Given this application of process mining, an institution might benefit from evaluating not just the bottlenecks but the particularly streamlined cases to determine of practice changes might be implemented to improve overall care.

Unfortunately, most medical conditions do not have well-defined, institutional guidelines from which to compare the quality of care. But perhaps this offers even more compelling potential for such unsupervised learning methods. Process mining could be used to gain early insights into patient management variability which could then be used to coordinate care, orders and resources as needed where delays or bottlenecks occur. Heterogeneously managed conditions that may currently lack guidelines, such as neonatal fever or even COVID-19, might in fact be ripe for identifying practice-based interventions which are most related to high-quality outcomes. Our process-mining approach could be used to identify transitions of care or patient movement through the hospital which makes nosocomial infections more likely. Alternatively, it could be used at the provider level to limit those transitions of care or exposure opportunities. Process mining might also be used to identify the most efficient triage or distribution of resources during a disaster when care is provided in an evolving context.

It is important to note that while allowing for these more unsupervised methods can help reveal in-vivo practice patterns, they may have unexpected results which require further investigation. For example, loops may develop in the pathway, which might seem difficult in the real-world. We noted a loop existed late in our stroke pathway where tPA was ordered and then PT/INR was ordered again (Figure 3). This initially seemed counter-intuitive, but further clinical review indicated that these orders are not part of the diagnostic and management process, but rather ordered for continued evaluation and monitoring after clot lysing medications are given. Thus, it is important to recognize that process mining is heavily reliant on the timing of an order and when it is expected to be carried out as in the case of those events which are not part of the primary diagnostic process but rather observation and monitoring. Similarly, process maps show overall average most common edge sequences, but their inherent variability means no single consistent interpretation can be drawn, as reflected by need to delete many sparse edges to allow for interpretability as well as the aforementioned loops. This is particularly confounding when the map includes the manual entry of test orders (like point-of-care tests) or back-dating verbal orders (which may occur for some acutely ill patients) which may result in confusion at minute-level resolution.

Furthermore, while we specifically chose to evaluate the acute management of stroke, an emergent condition which had a consensus pathway from which to compare our automatically generated map, the very existence of a consensus policy within our institution presents a possible catch-22. Process mining techniques may only serve to identify the care which has already been predicted to be most efficacious or which has been clearly defined with minimal room for variation. However, it is in these cases where the process is already known that make our mixed method approach, utilizing conformity and variation, more poignant, as discussed above.

While this study was performed at a single institution, limiting the broader generalizability of our particular graphical and conformity results given differences in practice pattern, data availability and labeling which might result in more or less interpretable results. However, this method should be replicable from data available from most other EHR’s with minimal effort required to identify how key events are stored and labeled.

## CONCLUSION

Process mining may be used not only to easily illustrate complex treatment pathways but to also identify process variability across encounters. We were able to automatically create a graphical representation for the care of acute stroke patients receiving tPA in our hospital by utilizing event log data captured in our EHR through process mining techniques. Notably, we were able to *qualitatively* map the order and timing of key events like admission, lab testing, CT scan ordering and delivery of tPA as well as visualize differences in these processes for fast and slow progression through the pathway. Our graphical results also allowed for *quantitative* evaluation of conformity at the encounter and cohort levels.

This mixed methods approach represents an essential data analysis step to improve complex care processes by automatically generating a map of them through process mining of existing event log data. It sets the stage for future work to identify high yield processes, bottlenecks, or where work happens in ways that deviate from best practice guidelines. Furthermore, its use may be broadened to those conditions which currently lack recommended processes to gain insights necessary to develop best practices.

## Data Availability

This research used data or services provided by STARR, Stanford medicine Research data Repository, a clinical data warehouse containing live Epic data from Stanford Health Care (SHC), the University Healthcare Alliance (UHA) and Packard Children's Health Alliance (PCHA) clinics and other auxiliary data from Hospital applications such as radiology PACS. The STARR platform is developed and operated by Stanford Medicine Research IT team and is made possible by Stanford School of Medicine Research Office.

## Acknowledgements

This research used data or services provided by STARR, Stanford medicine Research data Repository,” a clinical data warehouse containing live Epic data from Stanford Health Care (SHC), the University Healthcare Alliance (UHA) and Packard Children’s Health Alliance (PCHA) clinics and other auxiliary data from Hospital applications such as radiology PACS. The STARR platform is developed and operated by Stanford Medicine Research IT team and is made possible by Stanford School of Medicine Research Office.

The content is solely the responsibility of the authors and does not necessarily represent the official views of the NIH or Stanford Healthcare.

## APPENDIX

**Figure 5:**
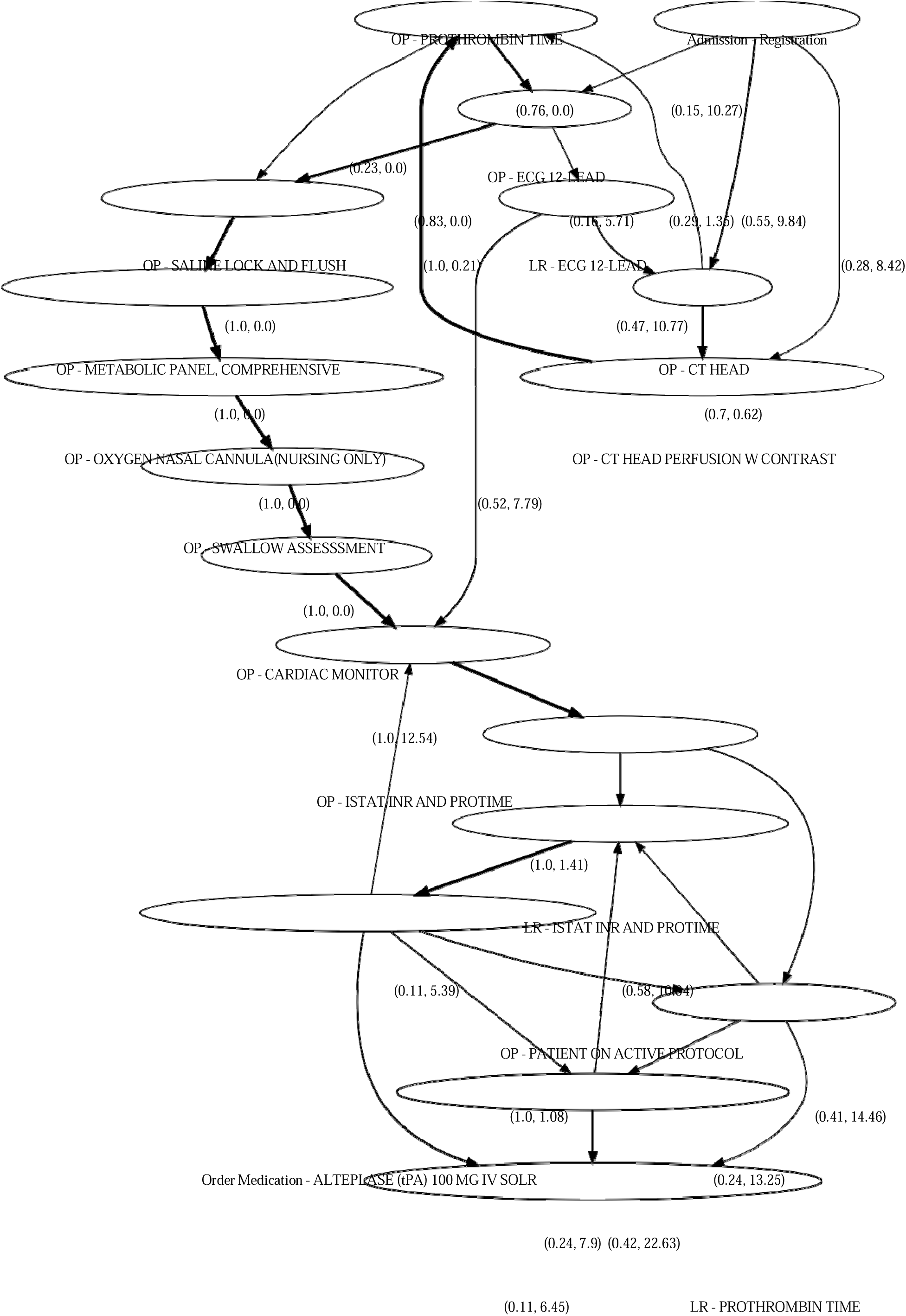
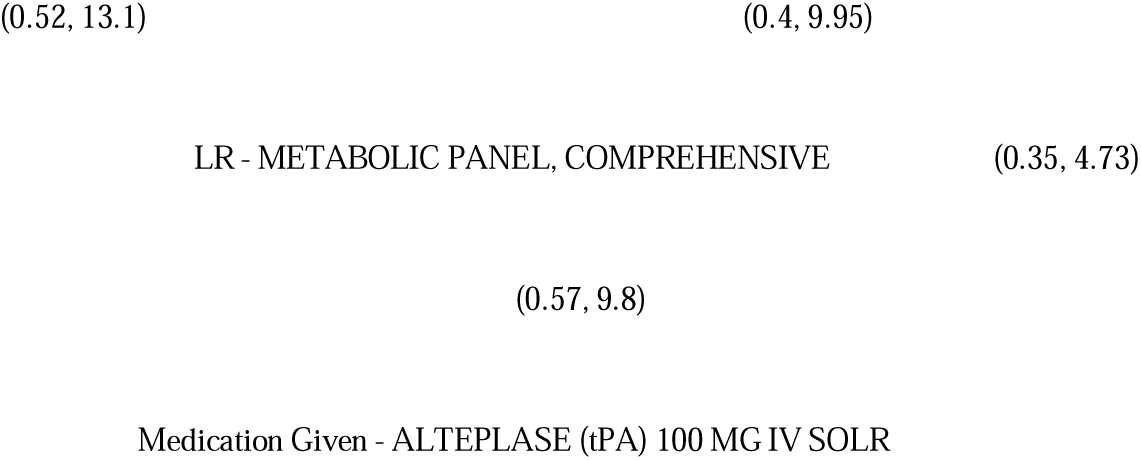
A sample process mining graph of the tPA-treated stroke patients, without applying the node clustering technique.

**Figure 6:**
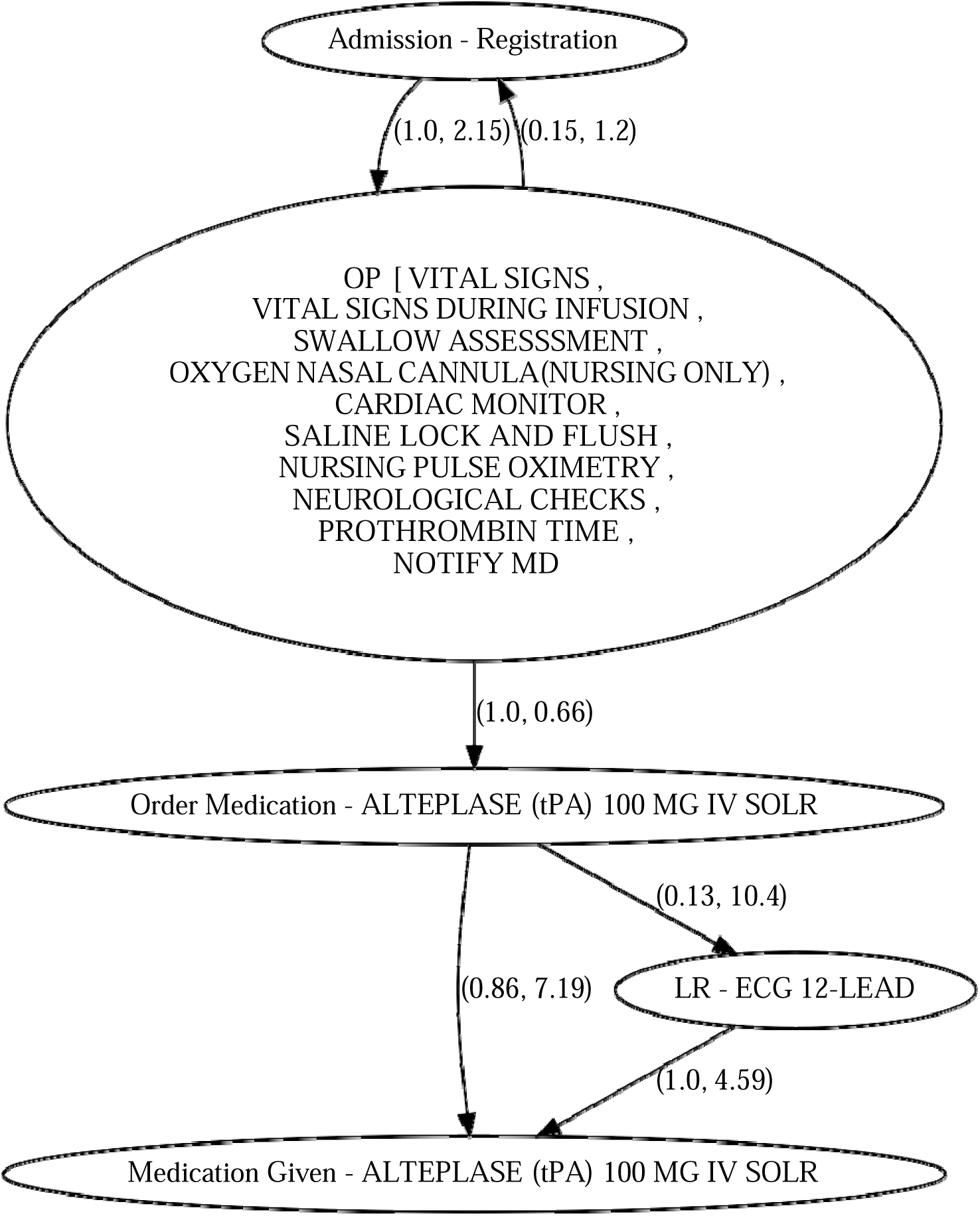
A sample process mining graph of the tPA-treated stroke patients with tPA administration with 25 min of their admission

